# Patterns of IgM Binding to Tumor Associated Antigen Peptides Correlate with the Type of Brain Tumors

**DOI:** 10.1101/2020.06.19.20135509

**Authors:** Dilyan Ferdinandov, Viktor Kostov, Maya Hadjieva, Velizar Shivarov, Assen Bussarsky, Anastas Pashov

## Abstract

The immune system can be used as a biosensor of the internal environment. Changes in the reactivities of the antibody repertoire can be used as a readout for a wide range of disturbances including various inflammatory conditions and malignant tumors. Extending our previous work based on IgM mimotope libraries, here we report our studies on the interpretability of profiles of IgM reactivities to a library of natural 15-mer peptides derived from 20 tumor associated antigens and 193 linear B cell epitopes involved in tumor pathogenesis. Sera from 21 patients with glioblastoma multiforme (GBM, n=10), brain metastases of other tumors (n=5) and non-tumor bearing neurosurgery patients (n=6) were used to probe their IgM reactivity with an array of 4526 peptide sequences. Using feature selection algorithms, we were able to extract profiles that separated well the three diagnostic groups with accuracy of up to 0.9. A key feature of the profiles extracted was their size (138 peptides for differentiating GBM and 340 – for tumor bearing patients) and origin from practically all tested antigens. Comparable numbers of reactivities were gained or lost in tumor bearing patients. A minimal set of the most significant 41 reactivities from 16 antigens contained disproportionately large number of epitopes from stromelysine-3 and erbB2 receptor with some of the reactivities gained and other lost in cancer patients. Epitopes from human papilloma virus 16 and HTLV-1 were included too. Some of the reactivities were readily interpretable both as antigen source and structural context (signal peptides). The interpretation of the rest requires further confirmatory studies. Thus, a set of natural peptides from tumor antigens readily provides profile of interpretable IgM reactivities which can serve as classifiers for clinically relevant patient stratification.

## Introduction

The malignant primary and metastatic brain tumors still have a poor prognosis and a high mortality rate. The early diagnosis, staging and monitoring are crucial for the effective treatment and for a possible increase of the survival rate [1].

In general, tumor biomarkers are products of damaged cells or pathogens and a certain concentration must be reached in order to detect them. Currently liquid biopsies are being used more and more for detection of molecular signatures of primary and metastatic cerebral neoplasms in blood or CSF [2]. “Liquid biopsies” refers to a number of minimally invasive methods and approaches of analyzing biomarkers circulating in body fluids in order to conduct a comprehensive profiling of a neoplasm for research or for further diagnostics.

The immune system detects cancer antigens. Appropriately reading its output could be an alternative source of clinical information [3; 4; 5]. Several studies have identified both whole proteins and peptides as IgG targeted antigens which are associated with brain tumors and provide useful clinical information [5; 6; 7; 8]. In all of these studies, the central paradigm is detecting IgG antibody responses to tumor associated antigens (TAA). Such responses exist in an apparent contradiction to some of the tenets of immune tolerance to self, since not all antibodies are to mutated epitopes.

There have been arguments in favor of quantitative self-tolerance thresholds [9]. Thus, the notion of self extends from qualitative to qualitative and quantitative characteristics of the antigen landscape. It could easily explain physiologically autoreactive antibodies binding to TAA in the absence of structural changes but simply as a result of the frequently observed overexpression of self-antigens in tumors. At the same time, quantitative self-tolerance is comprehensible mostly in terms of physiologically maintained titers of natural antibodies. This antibody compartment is also of limited inducibility mostly by danger signals rather than specific stimulation [10; 11]. Such properties make interpretations in the mainstream immunological context too hard so the IgM compartment is generally neglected. Yet its involvement in immune surveillance has been demonstrated [12; 13].

Previously, we have argued that the repertoire of IgM may be an untapped source of biomarkers [14; 15]. It was shown that mimotope libraries may serve as a source of probes to construct diagnostic profiles of IgM reactivities [14; 16]. As a proof or principle, such profiles were found to differentiate glioblastoma patients from patients with brain metastases or non-tumor patients. Although peptide mimotopes are structurally interpretable, the interpretation may be difficult when they represent conformational epitopes. To check if peptides from known TAA yield a more interpretable array of probes, here we repeat the same experimental setting with a commercial planar tumor antigen peptide microarray (PEPperPRINT®, Heidelberg). It contains a sequence set derived from 20 human proteins known as TAA and 193 linear B cell epitopes related to tumor pathology (PEPperPRINT®, Heidelberg). It was possible to identify sets of peptide reactivities that reproducibly differentiate patients with glioblastoma from those with brain metastases or non-tumor bearing. For most of the TAA identified like stromelysin-3, these results constitute plausible hypotheses for further exploration both of the TAA involvement in the tumor pathology as well as in the tumor immune system interface. Nevertheless, some of the results like HTLV1 epitopes may be explained better by cross-reactivity with unknown conformational epitopes.

## Materials and Methods

### Patients’ Sera

The presented pilot study includes surgically treated patients in the Clinic of Neurosurgery at St. Ivan Rilski University Hospital, Sofia, Bulgaria. All blood serum and brain tumor samples were obtained and analyzed after signed informed consent approved by the Ethics Committee at Medical University – Sofia. Only patients with proven glioblastoma or cerebral metastases of known origin were included. Patients who had undergone previous radio-, chemo- or immunotherapy, who suffered from an autoimmune disease or have had an inflammatory disease in the past three months were excluded.

Sera were obtained from randomly selected patients with glioblastoma multiforme (GBM, n=10), brain metastases of breast or lung cancers (meta, n=5), and non-tumor-bearing patients (contr, n= 6) with herniated disc surgery, trauma, etc. The sera were aliquoted and stored at − 20°C. Before staining, the sera were thawed; incubated for 30min at 37°C for dissolution of IgM complexes; diluted 1:100 with phosphate-buffered saline (PBS), pH 7.4, and 0.05% Tween 20 with 0.1% bovine serum albumin (BSA); further incubated for 30min at 37°C; and filtered through 0.22-μm filters before use.

### Peptide Microarray

Standard tumor antigen microarray chips PEPperCHIP® containing 4526 peptides produced by PEPperPRINT™ (Heidelberg, Germany) were used. The peptides are synthesized in situ attached to the surface through their C-terminus and a common spacer GGGS and are included in the microarray layout in duplicates. The majority of the peptides represent a scan of the sequences of 20 tumor associated antigens using a window of 15 amino acid residues shifted by two residues (Table 1). In addition, the peptide set contains also 193 B cell epitopes extracted from the Immune Epitope Database (IEDB, [17]) tagged “cancer” or “tumor”. This search strategy yielded a number of viral epitopes since the viruses themselves were implicated in cancer pathology.

**Table 1.**
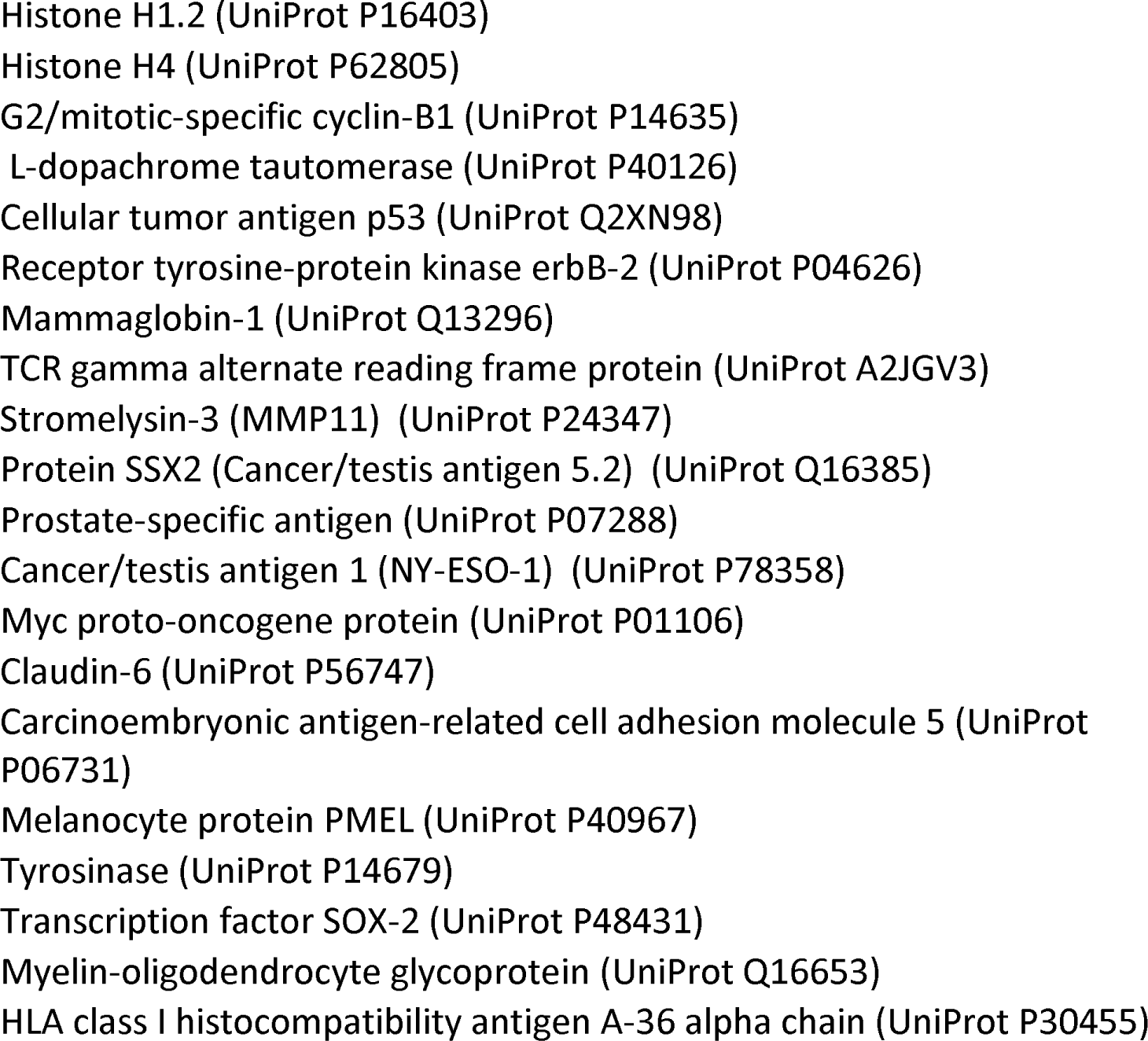
Tumor antigens included in the microarray.

The chips were blocked for 60min using PBS, pH 7.4, and 0.05% Tween 20 with 1% BSA on a rocker; washed 3 × 1min with PBS, pH 7.4, and 0.05% Tween 20; and incubated with sera in dilutions equivalent to 0.01 mg/ml IgM (∼1:100 serum dilution) on a rocker overnight at 4C. After 3 × 1-min washing, the chips were incubated with secondary antibodies at room temperature (RT), washed, rinsed with distilled water, and dried by spinning in a vertical position in empty 50-ml test tubes at 100 × g for 2 min.

### Microarray Data Analysis

The microarray images were acquired using a GenePix 4000 Microarray Scanner (Molecular Devices, USA). The densitometry was done using the GenePix R Pro v6.0 software. All further analysis was performed using publicly available packages of the R statistical environment for Windows (v3.4.1) (Bioconductor; Biostrings, limma, pepStat, sva, e1071, Rtsne, clvalid, etc.) as well as in-house developed R scripts (https://github.com/ansts/TAAIgM). The details of the analysis procedures are described elsewhere [14] with a few modifications.

The features selection algorithm consisted of a filter and a wrapper stage. In the filter stage:

1. All reactivities are represented as z-scores relative to the mean of the negative peptides (z0) and converted to false discovery rate adjusted p values. Retained are only peptides which display reactivities significantly higher than the background in more than 10 patients (at p<0.001 n=4285/4524).
2. For each peptide each patient’s serum reactivity is expressed as the greater absolute value of the 2 z-scores relative to the distributions of reactivities for the same peptide in the two alternative diagnostic groups (e.g. –a reactivity with a patient with GBM was tested against the mean reactivities of the patients with metastases or the non-tumor-bearing ones). Finally, each peptide is represented by the mean across patients of these z-scores - z1.
3. The intersection of the peptides found in (1.) and those for which z1>0.85 (feature set F1, n=378, used for the recursive elimination step) or z1>0.67 (feature set F2, n=963, used for the forward stepwise selection step, F2 includes F1). These cut offs allowed for the inclusion of the reactivities of significant intensity in more than half of the patients and with high discriminatory power with respect to the tested diagnostic groups.

The two sets of peptides were used further for the wrapper stage of feature selection which applied stepwise elimination or addition of features guided by a complex clustering criterion:

1. The selected features were further subjected to five cycles of recursive feature selection with backward (elimination, based on the F1 set) and forward step wise selection (based on the F2 set). The second step was used to add features to the set determined in the first step by screening a larger pool of second tier features. In both steps he criterion for selection is based on criteria for dichotomous clustering of the patients in the different diagnostic groups (GBM vs Control plus Metastatic or Control vs tumor bearing). The criterion was composed based on 3 internal clustering indices: Baker-Hubert gamma, Dunn and connectivity (see for details the supplemental methods of [14]). The clustering criteria had a stochastic term added which helps explore a larger feature space by repeating the procedure fivefold. Only the consensus reactivities from the 5 cycles are retained.
2. The recursive feature selection algorithm described was repeated in a “leave one out” scheme. Although the accuracy achieved exceeded 75%, the results were used further as a bootstrap exploration to estimate the variability of the feature sets in order to improve the capacity of the resultant classifiers to generalize. Only the reactivities recurrent in at least 50% of the bootstrap sets were retained and pooled to represent the final feature set of the classifier (see for details the supplemental methods of [14]).

In a third stage, the feature sets derived above were once again restricted based on the significance of the intensity of the reactivities relative to the means of the alternative diagnostic groups measured using the limma package from the Bioconductor set of R tools.

## Results

The study includes 21 patients divided into three groups according to the diagnosis: histologically proven glioblastoma (GBM, n=10), cerebral metastases (n=5, 4 lung cancer, 1 prostate cancer) or control (n=6). Control subjects were selected randomly out of cases with degenerative spine disease. After staining, reading, cleaning and normalizing the data represented the IgM reactivities of 21 patients with 4524 peptides from 20 tumor antigens and 193 known B cell epitopes of self or viral antigens related to cancer. The great majority of the peptides showed significant reactivity with the sera with 2974 being significant with each patient’s IgM and 4285 - with more than 10 patients. Due to the highly variable expression of different reactivities between the individuals, only a couple of peptides showed significantly different reactivity between diagnostic groups.

The patterns of reactivities (or using the machine learning parlance - features) of the IgM repertoire have an extreme interindividual variability [14]. They are affected by a great number of factors and, although apparently rich in information about the immunological status of each patient, the patterns are divergent and probably dynamic. Only combining them inclusively in patterns can lead to classifiers with a sufficient power. To this end, a battery of classical feature selection algorithms was applied. First the number of significant reactivities for each peptide and the difference from the mean reactivity of the alternative diagnoses for each patient were used to filter out the features with lower discriminatory power. This filtering stage was followed by the so-called wrapper algorithms. They test the capacity of the feature sets to produce classifiers that separate the diagnoses of interest in a measurable way and repeatedly replace features until the separation criterion reaches a maximum (see Materials and Methods for details). In the process, we incorporated a “leave one out” scheme to measure the degree of generalization of the prediction. The feature selection was performed on all sets of 20 out of 21 patients and at each step the resultant reactivity profile was used to predict the case left out. This involved teaching at each step a different optimized support vector machine model. The results are illustrated by two sets of figures each representing a 2-dimensional map of the cases (Supplemental material figures 1-42, the circled case is being predicted, the color background denotes the probability for the classification as GBM, resp.-non-tumor bearing Control). The map – a multidimensional scaling, is a 2-D representation preserving as best as possible the distances between the cases in the original n-dimensional space. Here n denotes the number of features in the profile. The figures (Supl. Figures 1-42) show the quality of the prediction in each of the “leave one out” steps. The overall performance of the feature selection step was good. The algorithm separating GBM cases from the rest yielded classifiers with accuracy 0.86 (Mathew’s correlation coefficient (MCC) - 0.72, F1 criterion – 0.83) and the one differentiating tumor bearing from non-tumor-bearing cases had accuracy 0.9 (MCC-0.77, F1 criterion - 0.84). Although these parameters were convincing, the diversity of the selected feature sets was too great. The sets differentiating GBM from the rest contained between 7 and 250 features while the one differentiating between tumor bearing and non-tumor bearing patients – between 431 and 525 features. Only 3 peptides were common for all GBM identifying patterns while 316 peptides were common for all feature sets predicting the presence of tumor.

As demonstrated previously [14], pooling the features that recur in at least 50% of the “leave one out” steps increases significantly the generalization (measured by MCC) of the resultant classifiers. Applying this approach in the present study yielded profiles of 138 peptides for GBM vs the rest and 340 peptides for Control vs Tumor Bearing classifiers. These sets performed sufficiently well separating the three diagnostic groups (Figures 1-4). While the classifiers were trained to separate dichotomously the GBM vs the rest, resp. - the Control vs tumor-bearing in both cases they separated reasonably well all three diagnostic groups (Figures 1 and 3). This is an indication that the peptide reactivities selected represent objective IgM reactivity patterns which readily differentiate the major diagnostic group studied. The unsupervised biclustering analysis finds comparably sized clusters of peptide IgM reactivities that are increased or decreased in the GBM patients with highly diverse individual patterns (Figure 2). The set differentiating tumor-bearing from non-tumor-bearing patients has a more complex composition (Figure 4). The two clusters of reactivities up or downregulated in tumor bearing patients are further subdivided in some that predominate in all tumor bearing and others that are found predominantly in patients with GBM or lung cancer metastasis.

**Figure 1.**
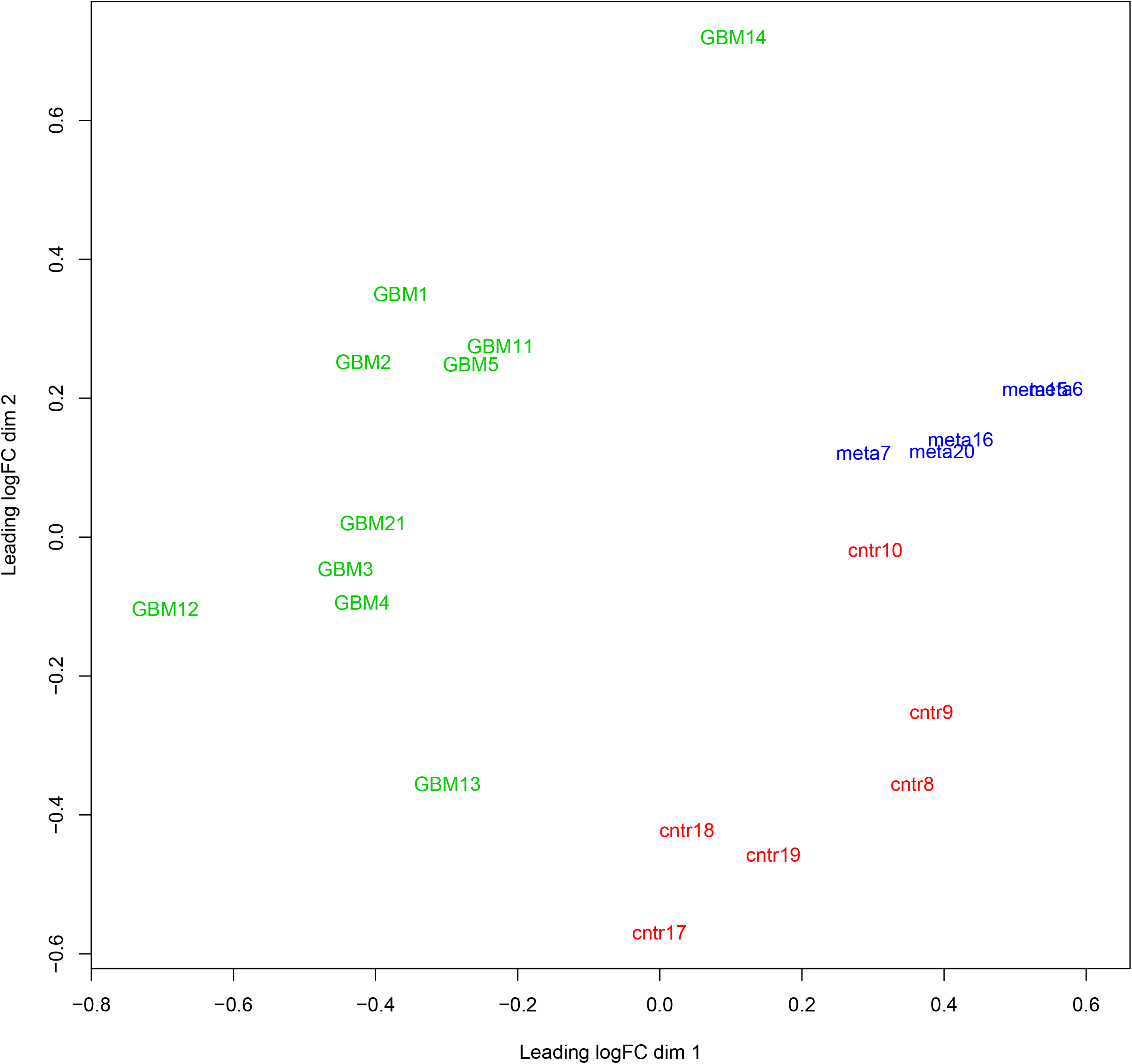
Multidimensional scaling projection of the patients’ serum IgM reactivity with peptide reactivities selected using a multistep feature selection algorithm which was previously shown to provide models that generalize well. The selected set of peptide reactivities separates perfectly the glioblastoma patients from the rest.

**Figure 2.**
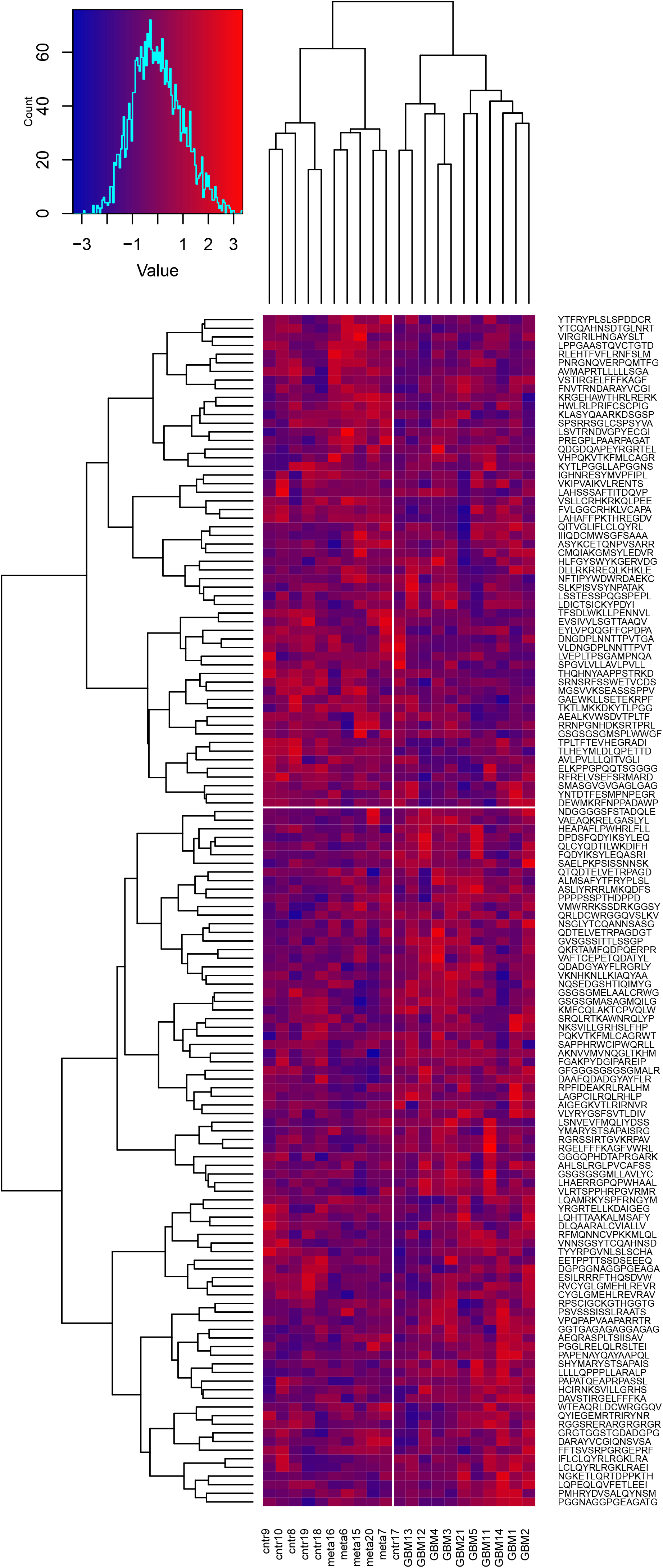
Biclustering analysis of the patients’ IgM reactivities with the optimal profile separating glioblastoma patients. The profile provides a good clustering of the non-tumor bearing patients.

**Figure 3.**
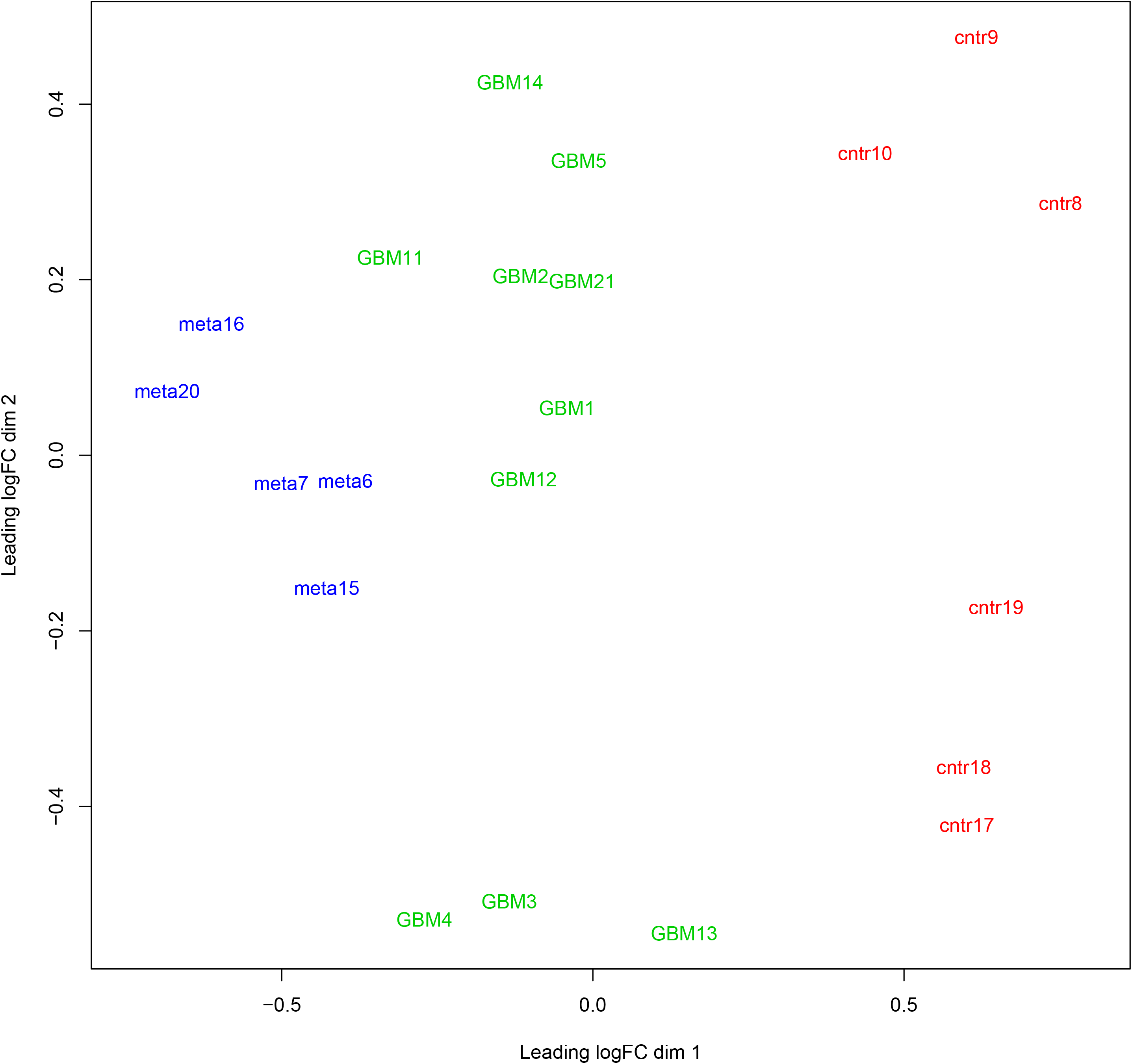
Multidimensional scaling projection of the patients’ serum IgM reactivity with peptide reactivities selected using a multistep feature selection algorithm. The selected set of peptide reactivities separates perfectly the non-tumor bearing from the cancer patients’ serum IgM reactivities but also GBM from metastases.

**Figure 4.**
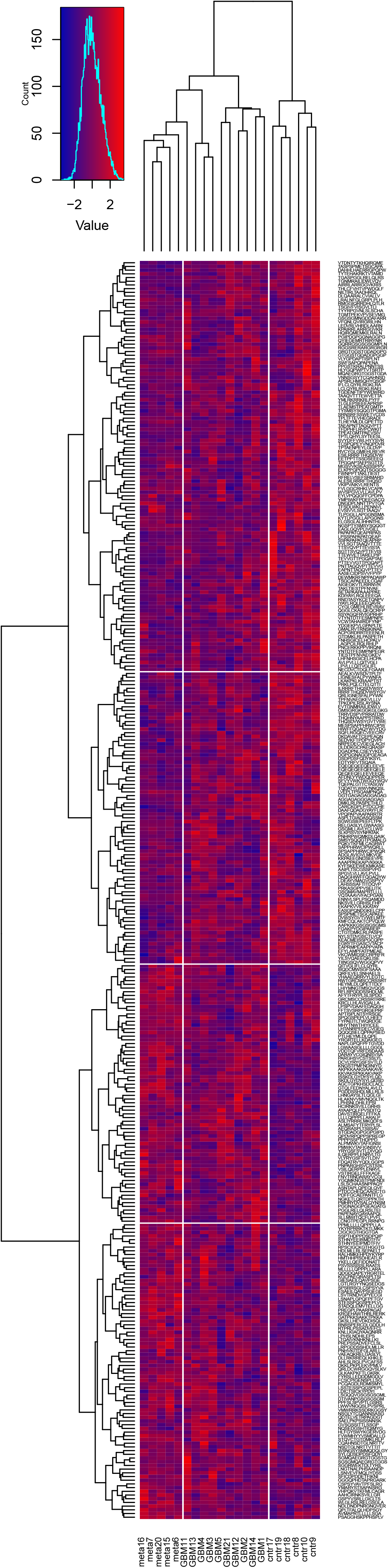
Biclustering analysis of the patients’ IgM reactivities with the optimal profile separating non-tumor bearing from cancer patients. The profile provides a good clustering not only of the non-tumor bearing patients but of all diagnostic groups. 190/340 reactivities are downregulated in at least some tumor-bearing patients.

The large number of peptide reactivities included in the patterns made the immunological interpretation of the reactivity to individual antigens and structures harder. Moreover, although the sets performed well, they featured considerable diversity of peptide reactivity patterns. In the selected feature sets no peptide reactivity differed significantly from the mean reactivities of the 2 alternative diagnostic groups at p<0.05 but 32 peptides form the GBM set trended towards a significant difference at p=0.051 and 19 from the Control set – at p<0.1. Next, an attempt was made to reduce the complexity of these patterns with minimal loss of information by focusing on these more significant reactivities. The union of the two sets yielded 44 reactivities but 3 peptide pairs represented adjacent sequence frames indicating a common shorter epitope. These pairs were replaced by the respective common epitope. The ultimate combined set contained 41 reactivities from 16 antigens (Table 2). This final minimal set proved superior in separating the three diagnostic groups (Figure 5) and clearly fell into 5 easily interpretable clusters (Figure 6): - cluster 1 – reactivities found predominantly in brain metastases, cluster 2 – reactivities characteristic of tumor-bearing patients, cluster 3 – reactivities in non-tumor-bearing patients which are usually lost in tumor bearing ones, cluster 4 - reactivities in non-tumor-bearing patients which are usually lost only in lung cancer metastases and cluster 5 – reactivities characteristic of GBM. It was interesting to find an abundance of epitopes from stromelysin-3(MMP11) and the receptor erbB-2. Another important observation was that reactivities to different epitopes from the same antigen had different behavior – some were up regulated and some were downregulated in tumor-bearing patients. Overall, a considerable number of reactivities tended to be lower in tumor-bearing patients.

**Table 2.**
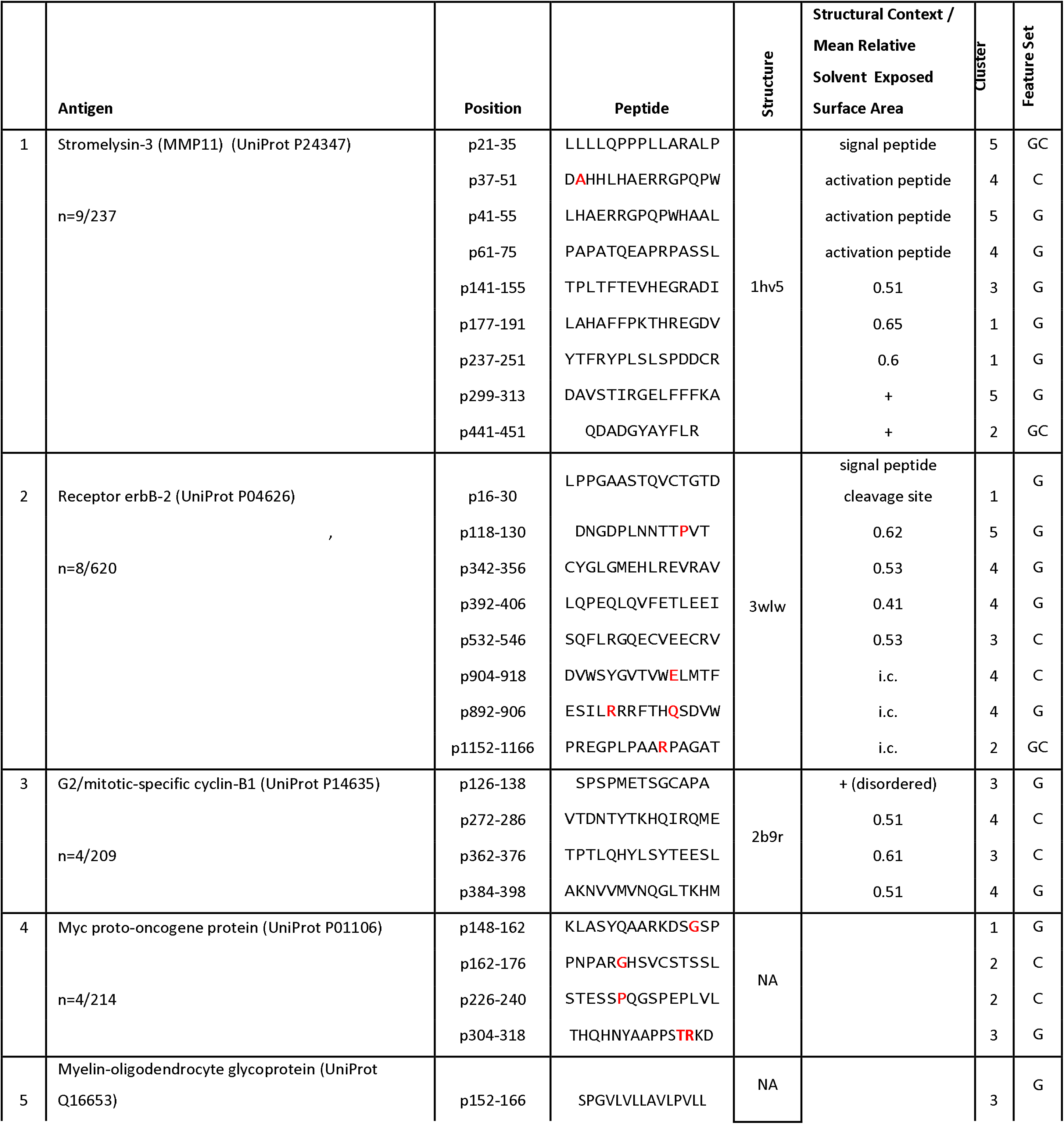
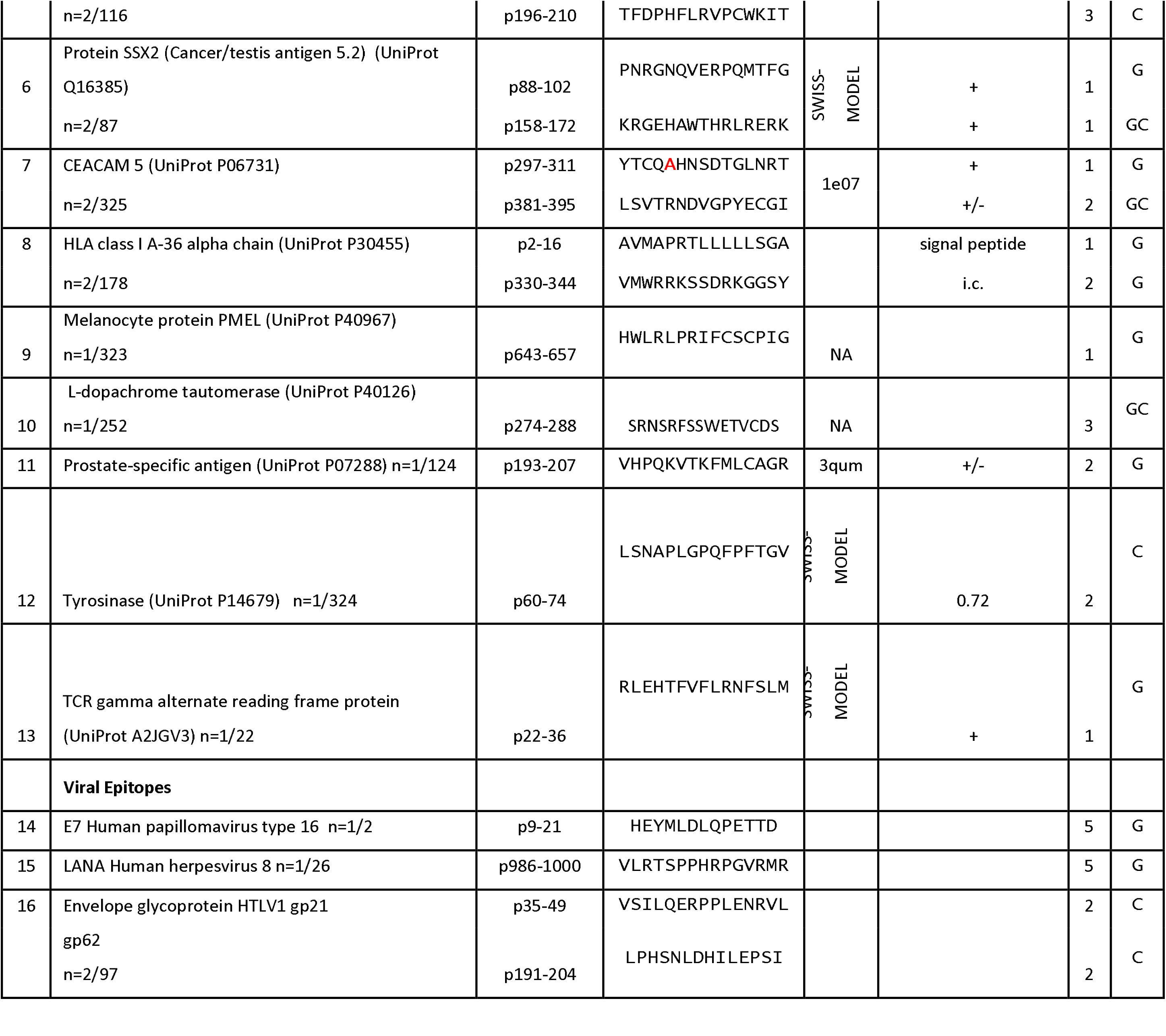
Antigen source and structural information for the minimal set of peptides representing a profile of IgM reactivities which separate the three diagnostic groups. In the Antigen column the numbers represent number of selected peptides relative to the number of peptides representing this antigen in the microarray. The red colored letters denote known and validated SNP variants. In the Structural Context column the figures represent the Mean Relative Solvent - Exposed Surface Area whenever it was possible to calculate it, “+”or “+/-” represent structural indications that the fragment might be exposed. For some peptides this column simply states their role as signal or activation peptides or that they are part of the intracellular portion for which there is no structure available. The column Cluster indicates the cluster (as shown on Figure 6) to which this peptide’s reactivity belongs: 1 – reactivities found predominantly in lung metastases, 2 – reactivities characteristic of tumor-bearing patients, 3 – reactivities in non-tumor-bearing patients which are usually lost in tumor bearing ones, 4 - reactivities in non-tumor-bearing patients which are usually lost only in lung cancer metastases and 5 – reactivities characteristic of GBM. The Feature Set column indicates the feature set the peptide belonged to after the wrapper method stage of the feature selection, G – GBM vs the rest, C – non-tumor-bearing vs the rest. GC stands for the peptides which were selected in both feature sets.

**Figure 5.**
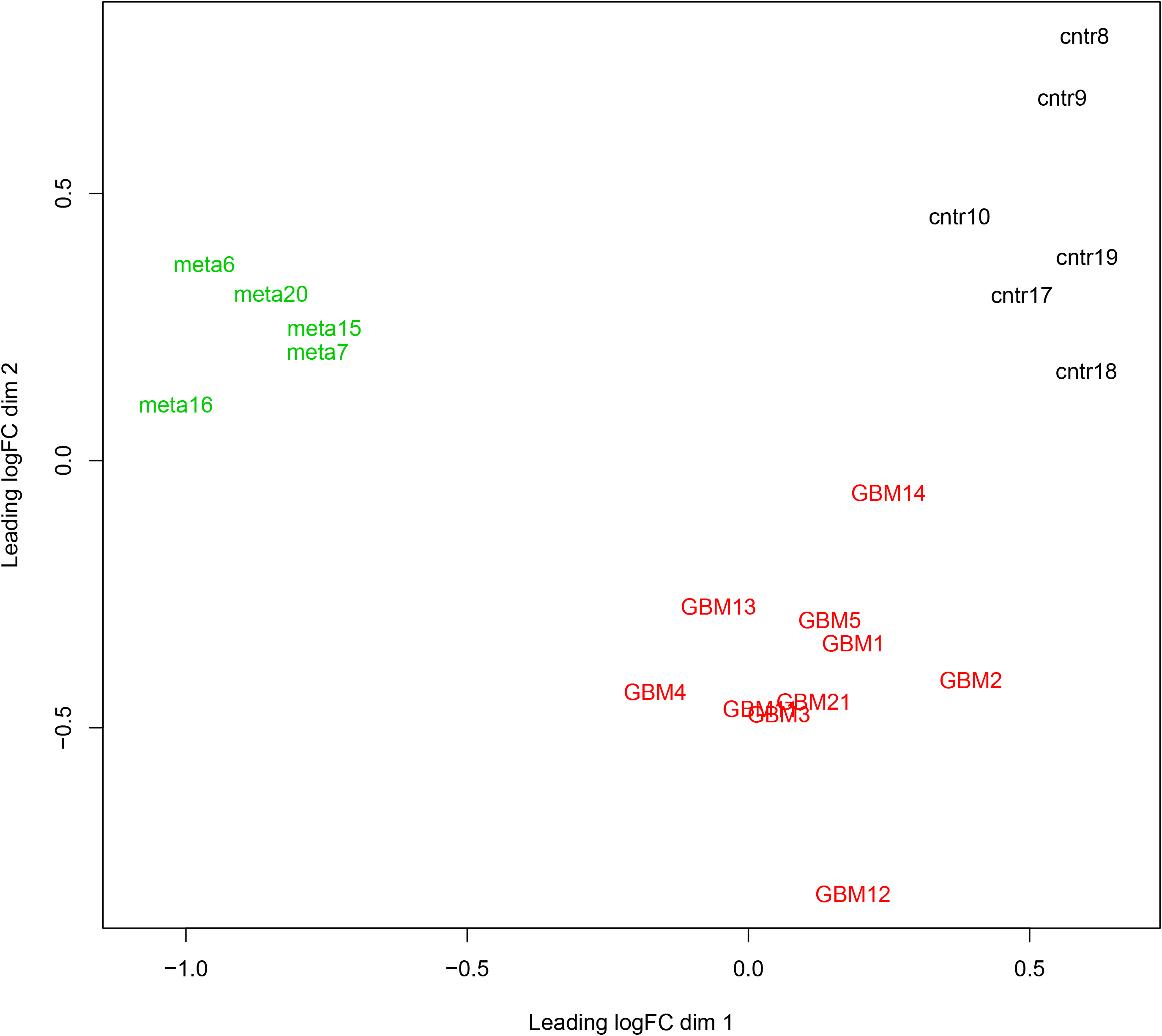
Multidimensional scaling projection of the patients’ serum IgM reactivity with a minimal set of 46/4524 peptide reactivities. The selected set of reactivities separates perfectly the three diagnostic groups.

**Figure 6.**
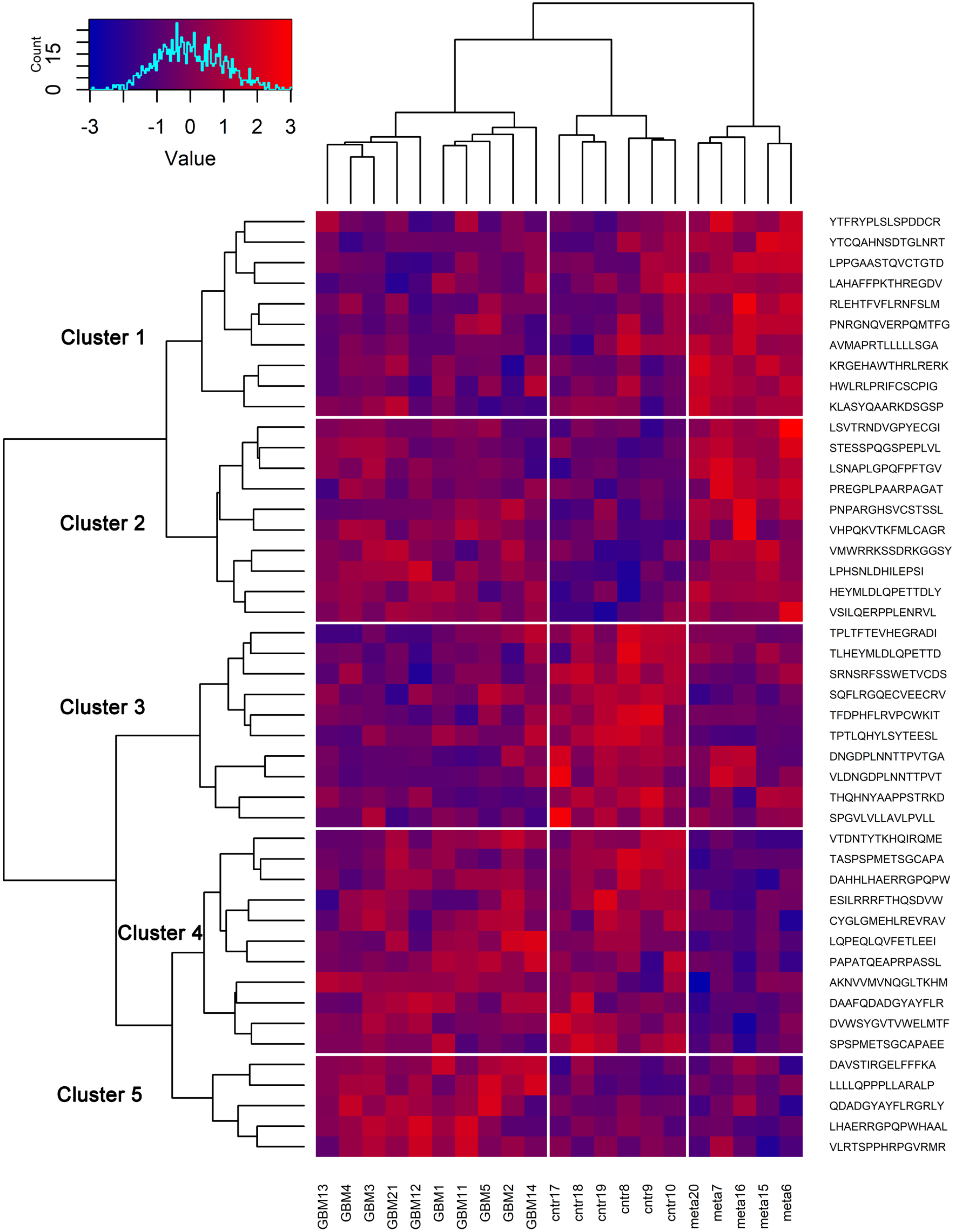
Biclustering analysis of the patients’ IgM reactivities with the minimal profile separating the three diagnostic groups. The profile provides a good clustering both of patients and of peptides which fall into 5 clusters: 1 – reactivities found predominantly in lung metastases, 2 – reactivities characteristic of tumor-bearing patients, 3 – reactivities in non-tumor-bearing patients which are usually lost in tumor bearing ones, 4 - reactivities in non-tumor-bearing patients which are usually lost only in lung cancer metastases and 5 – reactivities characteristic of GBM.

The distribution of the peptides in the minimal set did not differ significantly from the overall distribution between antigens (Chi square test, p<0.05) but the standardized residuals of the Chi square distribution corresponded to a significantly high number of peptides from stromelysin-3 (MMP-11) and from E7 of the human papilloma virus 16. Thus, there was a tendency to find many different potential epitopes from these antigens which stresses the significance of only the multiplicity but not of the occurrence. There was no clear correlation between the cluster number (the classification of the peptides with respect to presence or absence in different diagnoses) and their accessibility to the immune system. Interestingly, 6 peptides were from signal or activation peptides and 5/6 were characteristic of tumor bearing patients. Also, it was not surprising to find reactivities to papilloma virus (at lower levels of intensity also reactivities to herpes viruses were found – data not shown) but the surprising finding is the reactivity with 2 HTLV-1 epitopes because this virus is not common for the Bulgarian population.

## Discussion

In the present study, the patterns of reactivity of IgM with peptides from known TAA or cancer related linear B cell epitopes were studied in brain tumor patients. A great diversity of patterns of reactivity in the individual cases was observed. It is conceivable that the immune reorganization due to the presence of a malignant tumor is significant but still the amount of altered reactivities was in the hundreds and spanned most of the tested TAA.

Although rarely, IgM reactivity has been used to extract diagnostic reactivity profiles. In mice IgM reactivity to a set of autoantigens provided more consistently tumor related patterns than IgG [4]. Stafford et al. (2016) demonstrated that IgM reactivity to random peptides were more consistent than IgG both inter individually and in time which was interpreted in favor of IgG as a richer source of information [18]. The opposite argument stresses the necessity antibody repertoire profiles to be also sufficiently consistent and independent of the individual’s immunological history to be sources of generalizing classifiers or predictors. Even just for IgM, we observed such a diversity of IgM reactivity that patients’ sera sets which differed by only 2/21 cases (in the “leave one out” validation) selected greatly different feature profiles for the same classification task. Form 439 features found in any of the 21 sets classifying patients with GBM vs the rest only 123 (28%) were common for 12 or more sets and only 3 (0.68%) were common for all sets. Previously, we found a similar variability in the diagnostic profiles when using a library of peptide IgM mimotopes [14]. Finding classifiers working well with the training set when using efficient feature selection algorithms is not so surprising. The space of subsets of more than 100 features out of 4524 is for all practical purposes infinite (exceeding greatly 10^400^). So is its capacity to produce false classifiers overfitting to the training set. What comes as somewhat unexpected is that the uncovered diverse sets of features would together yield accuracy of classification in a leave one out validation of 0.86 and 0.9 even with so small training sets. Although diverse, these classifiers converge on the same decision. A possible explanation would be that the IgM repertoire readily provides efficient classifiers which converge on a common public repertoire profile while each patient exhibits at any given time just a subset of it. In other words, the data suggest a number of partially overlapping “repertoire phenotypes” exist among individuals. Some of these “repertoire phenotypes” may be actually temporary states of the repertoire which oscillate in time [19]. At any point of time, combinations of increasing numbers of “phenotypes” would yield classifiers asymptotically approximating the global public profile. Still, even those composite profiles would be different for the most part. Such a scenario would provide also opportunity for optimizing the “recombinations” of these profiles in order to achieve better generalization in the population. Previously, it was demonstrated that, in most cases, neither the features that recur in all profiles nor the simple sum of all features found in any profile form a working classifier [14]. It is rather the combination of features found in at least half of the “phenotypes” that work best. Probably, in the proposed profile recombination a minimal length of each profile needs to be present for the efficient combination of recurring features.

The key problem addressed in the present study was to provide data that can help test the interpretability of the IgM reactivity profiles. Namely, do actual peptides from known TAA and linear B cell epitopes provide more information when compared to arrays of random peptides [5] and synthetic mimotopes from phage display [14]? The extended profiles (Figures 1-4) contained peptides from most of the used antigens while the small optimized set still contained peptides from 13/20 tested proteins (Table 2) which show no significant difference in the overall distribution of reactivities among the tested proteins. On the other hand, the standardized residuals of the χ^2^ distribution yielded two proteins with excessive representation of peptides - stromelysin-3(MMP11) and the receptor erbB-2.

Finding that IgM reactivities to these two TAA help classify brain tumors is a plausible piece of information which should be explored further. All tumor antigens represented in the extended profiles also make sense to some extend except for PSA (tissue kallikrein 3, KLK3). KLK6 and KLK8 (absent from the tested TAA set) are expressed in brain tissues and the respective peptide shares some homology with KLK3. The antigen set included in the microarray does not represent other kallikreins as well as many other self-antigens so all reactivities should be interpreted in view of the possible polyspecificity of the IgM antibodies and the high probability of cross-reactivity.

What is more confusing is the distribution of the peptides coming from the same TAA with respect to the expression in the different diagnostic groups. Even for antigens that are overrepresented like stromelysin-3 and erbB2, reactivities from the same antigen can fall in all possible groups – some correlating with different brain tumors, some with their absence. The major mechanism of changes in the IgM repertoire may be fluctuations caused by inflammatory signals, fixation of natural antibodies in the tissues or changes in the composition of the repertoire of the respective plasma cells in the bone marrow and much less the classical immune response. These characteristics may explain the great number of reactivities found in non-tumor bearing patients and lost in those with tumors.

The IgM repertoire consists to a great extent of inherently polyspecific clones and most of the bound structures are expected to be conformational epitopes. These two facts should be reflected in the reactivities to the 15-me epitopes of known TAA tested. While some of the observed reactivities may be actual autoreactivities involved in tumor surveillance others may be simply cross-reactivities with little significance in vivo. The concentration of reactivities in TAA associated with brain tumors like stromelysin-3 [20] and erbB2 [21; 22] indicates, nevertheless, an intense interaction of these TAA with the IgM repertoire in the pathogenesis of the tumors.

Interestingly, the best classifier contained 6 reactivities to signal peptides and 5 of them were characteristic of tumor bearing patients. Signal peptides have been described as tumor specific epitopes before[23; 24]. Similarly logical seems the finding of reactivities to papillomavirus 16 exclusively in GBM patients which confirms a previous report of association of this virus with glioblastoma [25; 26]. On the other hand, the intriguing finding of reactivities to HTLV-1 seems more probably due to cross-reactivities because repeated testing of Bulgarian blood donors has failed to find HTLV-1 although there are reported endemic HTLV-1 infection in neighboring Rumania ([27] and Prof. Rada Argirova, personal communication).

Thus, the proposed IgM repertoire based diagnostic classifiers have the potential to provide a fast, non-invasive and inexpensive tool for early and accurate diagnosis of malignant tumors affecting the nervous system. By using natural peptides from known tumor associated antigens, it is possible also to find interpretable reactivities pointing to known or unknown prognostic markers which would give new opportunities in the treatment of the discussed diseases.

## Data Availability

All data and scripts are available from GitHub.

https://github.com/ansts/TAAIgM

## Acknowledgements

This work was performed with the support of intramural grant of the Medical University in Sofia 355/2015 and the Bulgarian Fund for Scientific Research Grant D01-11/2016.

